# Consultation informs strategies to improve functional evidence use in variant classification

**DOI:** 10.1101/2024.12.04.24318523

**Authors:** Rehan M. Villani, Bronwyn Terrill, Emma Tudini, Maddison E. McKenzie, Corrina C. Cliffe, Christopher N. Hahn, Ben Lundie, Tessa Mattiske, Ebony Matotek, Abbye E. McEwen, Sarah L. Nickerson, James Breen, Douglas M. Fowler, John Christodoulou, Lea Starita, Alan F. Rubin, Amanda B. Spurdle

**Affiliations:** QIMR Berghofer Medical Research Institute, Brisbane, Queensland, Australia; Australian Genomics, Murdoch Children’s Research Institute, Melbourne, Australia; Clinical Translation and Engagement Platform, Garvan Institute of Medical Research, Sydney, Australia; Faculty of Medicine and Health, UNSW Sydney, Sydney, Australia; Douglass Hanly Moir Pathology, Macquarie Park, Australia; Department of Genetics and Molecular Pathology, SA Pathology, Adelaide, Australia; Pathology Queensland, Queensland Health, Brisbane, Australia; University of Queensland, Brisbane, Australia; Australian Genomics, Australian Functional Genomics Network; Murdoch Childrens Research Institute, Parkville, Australia; Department of Laboratory Medicine and Pathology, University of Washington, Seattle, United States; Department of Genome Sciences, University of Washington, Seattle, United States; Brotman Baty Institute, Seattle, United States; Department of Diagnostic Genomics, PathWest, QEII Medical Centre, Perth, Australia; Black Ochre Data Labs (Indigenous Genomics), Telethon Kids Institute & Australian National University, Adelaide, Australia; Department of Bioengineering, University of Washington, Seattle, United States; Genomic Medicine Theme, Murdoch Children’s Research Institute, Melbourne, Australia; Department of Paediatrics, University of Melbourne, Melbourne, Australia; The Walter and Eliza Hall Institute of Medical Research, University of Melbourne, Melbourne, Australia; Department of Medical Biology, University of Melbourne, Melbourne, Australia

**Keywords:** Functional evidence, variant classification, diagnostic genetics, education, assay

## Abstract

To determine if a variant identified by diagnostic genetic testing is causal for disease, applied genetics professionals evaluate all available evidence to assign a clinical classification. Experimental assay data can provide strong functional evidence for or against pathogenicity in variant classification, but appears to be underutilised. We surveyed genetic diagnostic professionals in Australasia to assess their application of functional evidence in clinical practice. Results indicated that survey respondents are not confident to apply functional evidence, mainly due to uncertainty around practice recommendations. Respondents also identified need for support resources, educational opportunities, and in particular requested expert recommendations and updated practice guidelines to improve translation of experimental data to curation evidence. As an initial step, we have collated a list of functional assays recommended by 19 ClinGen Variant Curation Expert Panels as a source of international expert opinion on functional evidence evaluation. Additional support resources for diagnostic practice are in development.

## Introduction

A genetic diagnosis clarifies the underlying molecular cause of an inherited condition, and underpins informed treatment, management and counselling for individuals with inherited disease. However, classifying the clinical significance of genetic variants remains challenging, and many individuals with suspected inherited disease do not receive a molecular diagnosis. This situation is unlikely to improve unless additional sources of evidence can be effectively included in the curation process.

Recommendations for the classification of genetic variants developed by the American College of Medical Genetics and Genomics (ACMG) and Association for Molecular Pathology (AMP), now used worldwide, dictate that experimental data from “well-established functional studies” may be used as functional evidence in variant classification (1). Subsequent ClinGen recommendations (2) provide further guidance for the application of functional assays. These recommendations suggest that functional assay results must be rigorously evaluated to determine if the assay is appropriate, and the strength of evidence applicable in curation must be determined. Subject to calibration, functional evidence can be applied with a level of strength that is sufficient to shift a variant classification from one tier to another (code PS3 or BS3, Strong level of evidence) (1).

Functional evidence is highly valuable for variant classification as it is not limited by access to information or material from an affected individual, and can even be generated prospectively, prior to observation of a variant in the clinical setting.

ClinGen Variant Curation Expert Panels (VCEPs) define gene-specific adaptations of ACMG/AMP guidelines that have been shown to improve classification compared to baseline guidelines (3). All approved VCEP guideline specifications are publicly available (4). A 2019 review of six ClinGen VCEPs showed discordance in functional evidence application in their specialised guidelines (5). Shariant, a variant curation sharing platform for Australian and New Zealand laboratories, identified that functional evidence was applied in only 10% of diagnostic variant classifications (6). Application of high-throughput assays is leading to a rapid increase in the availability and generation of functional data (7), and resources such as MaveDB (8) are making datasets increasingly accessible for clinical and research use (9, 10). Given this rapid rise in generation of functional assay data, increasing the application of functional evidence is critical to improve diagnostic yield.

We undertook a study to investigate Australasian genomic diagnostics professionals’ perceptions and experience of evaluating functional evidence in diagnostic curation, and their education and support needs to improve application of this evidence type. We also reviewed and summarized current expert guidance documents from 34 released ClinGen VCEP Criteria Specifications, to determine their value as a practice resource for functional evidence application.

## Materials (or Subjects) and Methods

### Survey

A needs assessment survey aligned with the Program Logic model for genomics education (11) was developed by an interdisciplinary team of genetics professionals, based on a rapid literature review. Survey data was collected online and managed using REDCap electronic data capture (12) hosted at QIMR Berghofer and results collected via csv/R data download. The survey was distributed by the Human Genetics Society of Australasia, the Australasian Society of Diagnostic Genomics, and additional national genomics networks including the Australian Functional Genomics Network tools (13) and the Shariant user group (6). Information collected included demographics, curation experience and practice, education and implementation needs and attitudes of the participants (Supplementary Information 1). The introductory text for the survey included definitions of terms, a description of the intended use of the survey results, and consent statements. This research study was approved by the Human Research Ethics Committee of QIMR Berghofer, Project ID P3920.

### VCEP functional evidence review

Specific functional assays within VCEP Criteria Specifications (CSpecs) as of 16 July 2024 were collated by: accessing each VCEP functional evidence criteria specification (PS3, BS3), and sequentially extracting all referenced assays to collect the PMID, gene assayed, evidence level recommended by the VCEP and any specific context given for the assay. The assay was then reviewed for the type of assay, number of variants covered (all variants, including unknown and those denoted as controls), along with additional descriptors regarding the assay class and features (Supplementary Table 1).

### Data analysis

R (R version 4.3.2 (2023-10-31 ucrt))(14) /RStudio (2024.4.2.764) (15) was used to perform analyses and generate figures. This included calculating summary statistics, such as means and confidence intervals, and the various counts provided within this report.

## Results

Thirty three (33) genomics diagnostic professionals responded to the survey, including 27 scientists (10 research and 17 clinical diagnostic) and 6 genetics clinicians (various specialties but all with genetics focus). All participants indicated involvement with variant curation, and 23 denoted that this included curation in a clinical setting. The participants reported a range of variant curation experience, with an average of 12 years (±9.37SD).

Participants varied in their confidence/comfort in evaluating functional assays across five main categories (Figure 1). Comfort with functional assay interpretation (survey options comfortable or very comfortable) was indicated for: half of participants (17/32) for animal models; fewer than half for biochemical assays (14/32), transcript assays (15/32) or cell models (12/31); and only 6/32 for high-throughput assays. There was also substantial variation in participant practice and awareness of current recommendations and resources to evaluate functional evidence. A large proportion of participants were unaware of current recommendations and tools, and fewer than a third of participants actively used the provided recommendations and available tools. In particular, very few participants used the ClinGen Sequence Variant Interpretation (SVI) Functional Assay Documentation Worksheet (3/33) (16) or the VCEP CSpecs (5/33) for functional evidence evaluation.

**Figure 1.**
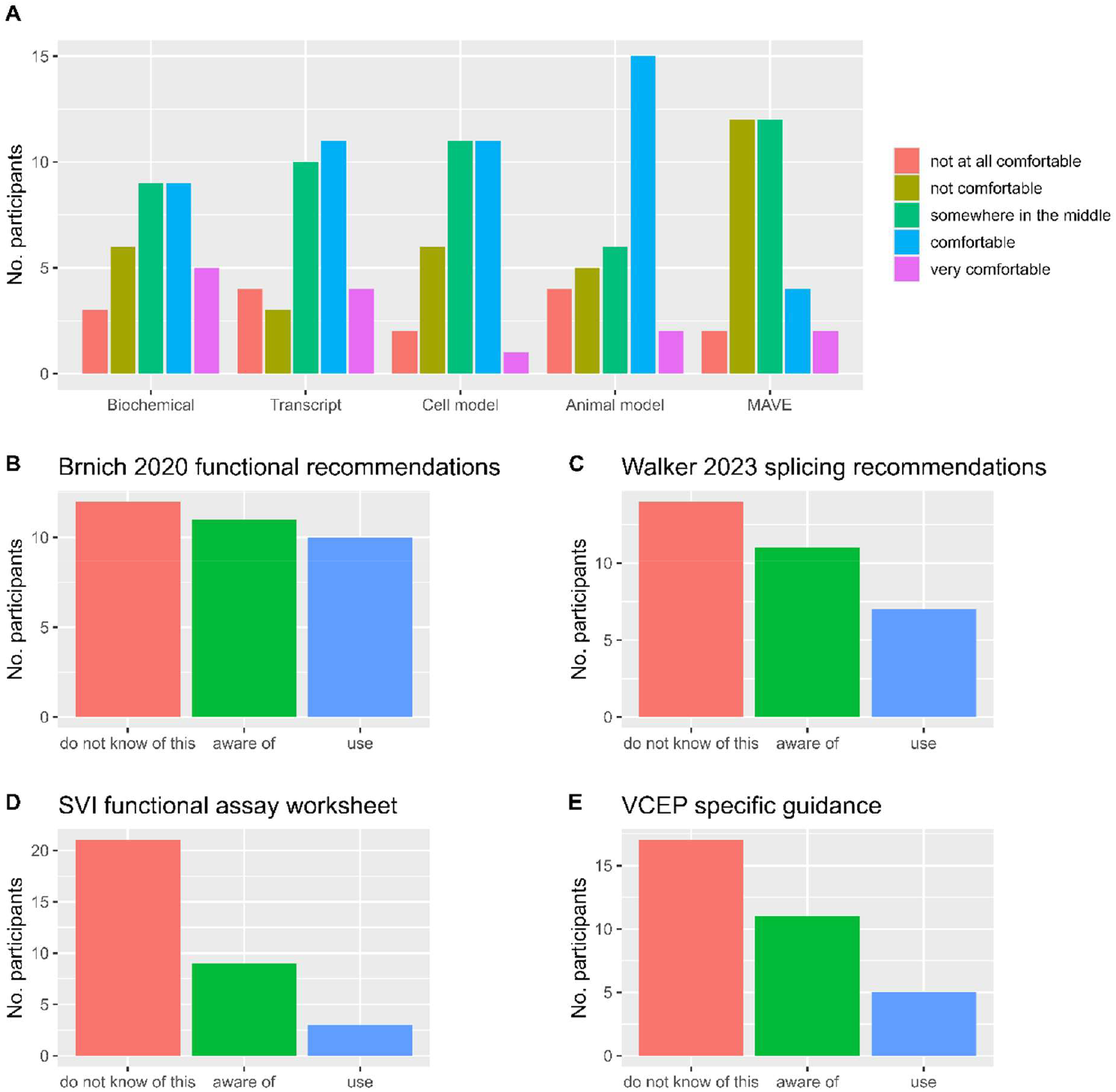
Comfort and familiarity of survey participants with the ACMG/AMP framework and associated recommendations and tools for functional evidence application. A) Confidence level of participants (n=33) according to the rating scale ‘1 = not at all comfortable’ to ‘5 = very comfortable’ for each of the indicated functional evidence types: biochemical assays (e.g. enzyme assays), transcript assays (e.g. splicing assays or transcriptome data), cell models (for e.g. *in vitro* cell assays), animal models (e.g. mouse models) and high throughput functional assays (including multiplexed assays of variant effect). B) – E) shows number of participants according to awareness level category (’use’, ‘are aware or’, or ‘do not know of’) for the indicated resources for functional evidence evaluation (33 participants each, unless otherwise noted). Note: awareness level was assessed independent of the standard operating procedure used by their practice/institution. B) Awareness of ClinGen Sequence Variant Interpretation (SVI) Working Group functional evidence recommendations (2). C) Awareness of ClinGen SVI Splicing Subgroup recommendations (20), n = 32 (one “Not Applicable” result excluded). D) Awareness of ClinGen SVI Functional Assay Documentation Worksheet (16). E) Awareness of VCEP specific guidance on functional evidence use (4).

Participants were asked to list barriers, enablers and training preferences for using functional evidence. The largest barriers to using functional evidence (Figure 2A) were ‘Lack of familiarity with a specific assay’ (20/33) and inability to ‘find functional data’ (19/33). Education (25/33), methods (23/33) and additional guidelines (22/33) were identified as potential enablers (Figure 2B). An overwhelming majority of participants (31/33) indicated that they ‘would be interested in training/further education and/or additional resources…’. The preferred educational strategies to increase/improve use and/or understanding (Figure 2C) were: expert guidance documents (24/33), closely followed by tools for evaluation (23/33), and online learning resources (23/33).

**Figure 2.**
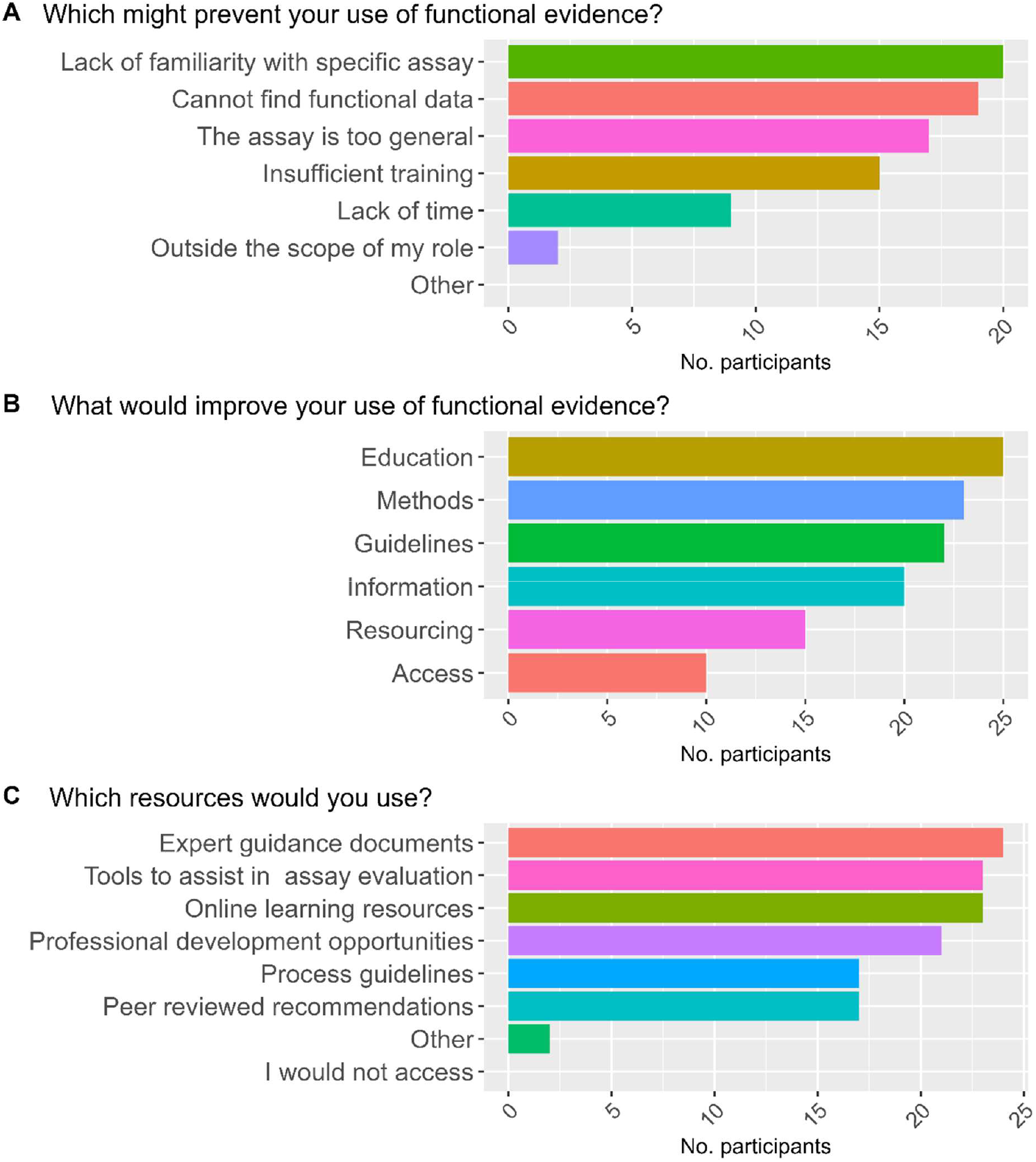
Participant views related to functional evidence application. Viewpoints were framed around three questions (A-C) on barriers, enablers and resources for future use, with option to select all that apply, and to provide free text answers to an “other” option. A) Which of the following might prevent your use of functional evidence in variant classification? B) What would improve your interaction with and use of functional evidence (existing or not)? C) Which of the following training and/or additional resources would you use to increase/improve your use/understanding of applying functional evidence?

We next undertook to review of a source of expert advice, specifications from approved ClinGen VCEPs, and extracted information on the specific assays mentioned. Of the 34 ClinGen VCEP CSpecs assessed, 19 VCEPs mentioned specific publications across 33 genes (Figure 3). Overall, 226 assays were specifically mentioned within VCEP CSpecs, with between 1 and 44 assays per gene (Figure 3A). Recommended assays covered more than 45,000 variants. Most assays were lower throughput (Figure 3B), with only 6 multiplexed assays of variant effect mentioned. VCEPs recommended application across all evidence strengths (Figure 3C and D). Additionally, the SVI functional working sheet was used in original or adapted form by only 10/34 VCEPs, suggesting that improved awareness of available resources will benefit practitioners broadly.

**Figure 3.**
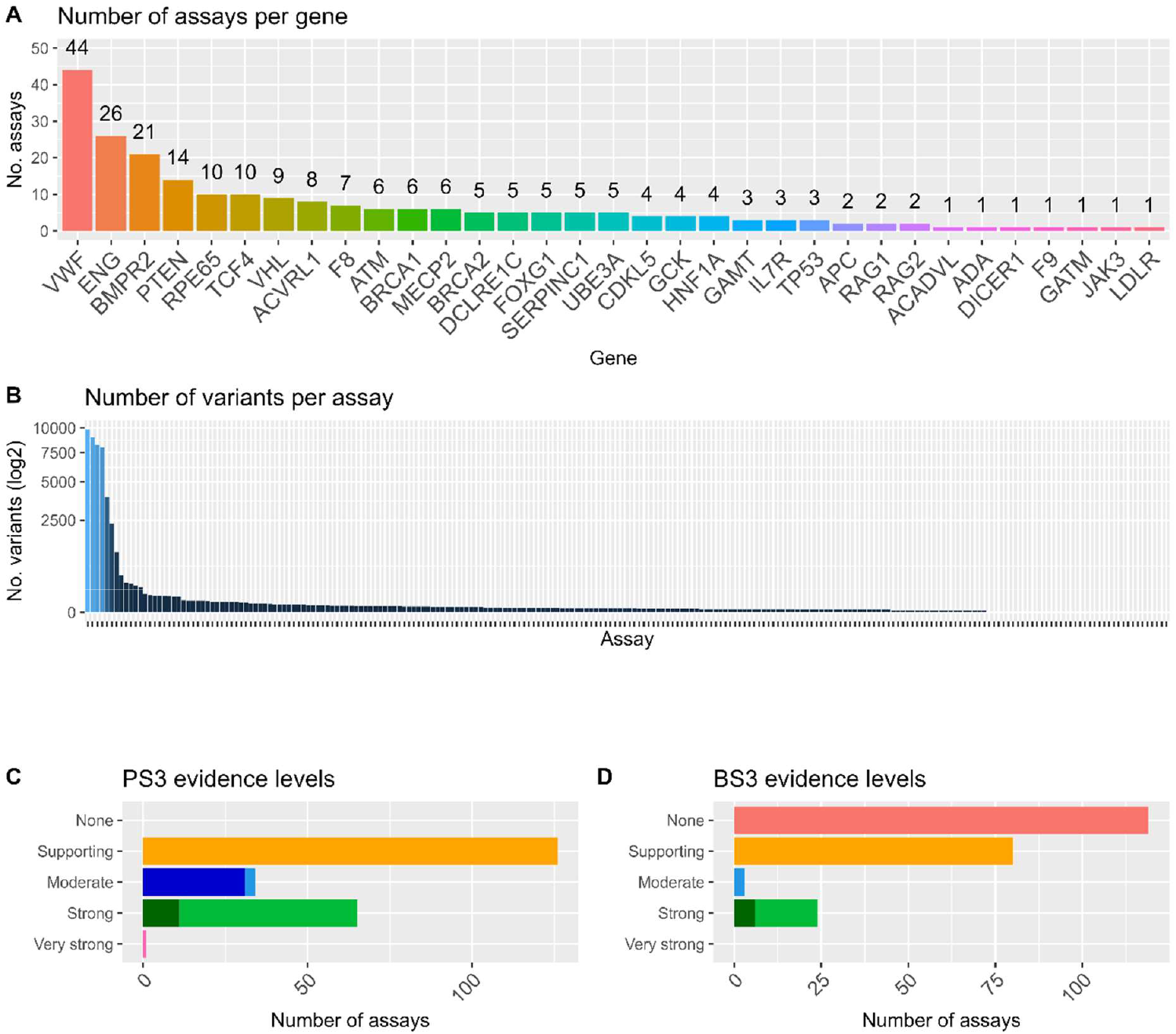
Summary of functional assays specifically mentioned within VCEP CSpecs. A) Number of assays identified for each of the 33 genes covered by the VCEP CSpecs (total n=226). B) Number of variants per assay across all VCEP CSpecs. C) Distribution of pathogenic evidence strength (applicable under ACMG/AMP code PS3) recommended across all the specific assays listed in CSpecs. D) Distribution of benign evidence strength (applicable under ACMG/AMP code BS3) recommended across all the specific assays listed in CSpecs. A full list of specific assays collated can be found in Supplementary Table 1. Darker bar regions indicate where VCEPs denote additional considerations and/or modifications for application of the given evidence level.

## Discussion

Improved functional evidence incorporation in variant classification is critical to enhance the benefits of genetic diagnostic testing. Our survey results indicate that functional evidence use may be limited because clinical genomics professionals lack confidence in functional data evaluation. Participants wanted expert guidance, tools and education to support practice. The majority of survey participants did not use, and a substantial proportion were unaware of, the currently available recommendations and resources - despite them indicating expert guidance and tools as the preferred method to improve functional evidence use. These results echo other areas of genomics implementation and education (11, 17, 18) and suggest a need for effective dissemination suited to the audience, and ideally evaluation of future tools and guidance to enable improved practice.

The survey findings demonstrate a need for increased availability and awareness of functional evidence advice and resources across all levels of curator expertise. Indeed, the majority (72.7%) of survey respondents indicated expert guidance documents would be useful. As an initial step to address the participant needs, we collated expert guidance relating to functional evidence application within an existing global resource: the ClinGen Variant Curation Expert Panel (VCEP) specifications (CSpecs). Our review of the current VCEP CSpecs assessed the type and extent of information available relating to functional assays, and whether they could be used as a source of expert advice on specific assays. This review revealed a source and wealth of international expert guidance on functional evidence application for selected high clinical impact genes, including identification of specific assays and applicable evidence weights.

Notably, participant comfort was lowest for application of high throughput assays, and relatively few were identified within VCEP-recommended assays. Given the increasing generation of high throughput data, and potential for clinical adoption (10, 19), our findings raise the importance of promoting international collaboration to share not only functional data (7, 8), but also resources and training approaches to enable use of these data in clinical curation. To this end, we provide our survey and VCEP assay collation for others to use in their local context. Our results highlight the value of expert guidance, tools and education and indicate the importance of an international campaign for collaborative practice development to maximise functional evidence application in clinical practice.

## Data Availability Statement

Weblinks used in study

SVI Functional Assay Documentation Worksheet; https://clinicalgenome.org/docs/svi-functional-assay-documentation-worksheet/

VCEPs; https://clinicalgenome.org/affiliation/vcep/#ep_table_heading

## Code Availability

The code and datasets generated during this study are available through github, https://github.com/ReeVee2006/fe_use

## Acknowledgment

We would like to thank the participants of the Functional Evidence Use survey, and acknowledge contribution of current and former members of the ClinGen VCEPs to development of VCEP specifications.

## Author Contribution Statement

Conceptualization: RMV, BT, CC, TM, AEM, AFR, ABS

Methodology: RMV, BT, TM, AEM, LS, AFR, ABS

Formal analysis: RMV, BT, ABS

Investigation: RV, MEM

Resources: RV, MEM

Data Curation: RV, MEM

Writing - Original Draft: RV, ABS

Writing - Review & Editing: RMV, BT, ET, CC, CNH, BL, TM, EPM, AEM, SLN, JB, DMF, JC, LS, AFR, ABS

Visualization: RV

Supervision: RV, ABS

Project administration: RV, AFR, ABS

Funding acquisition: DMF, AFR, ABS

## Ethical Approval

The ethical aspects of this research project were approved by the Human Research Ethics Committee of QIMR Berghofer, Project ID P3920.

