## Supplemental information 1 for "Consultation informs strategies to improve functional evidence use in variant classification"

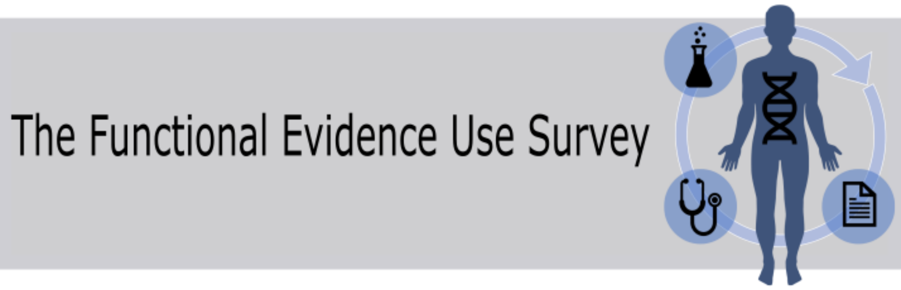


Returning?

A A A

**The Functional Evidence Use Survey**

**Understanding experimental data use in clinical genomic testing**

Multiplexed Assays of Variant Effects, or MAVEs, are large experimental assays that create functional evidence for many genetic variants, see the MaveDB. This survey aims to investigate the current use of functional (experimental) data, including MAVEs, and application of functional evidence for clinical variant classification. Results from this survey will inform the MAVE Education Project, which aims to increase the use of functional evidence in variant classification in the Australian clinical diagnostic setting.

The MAVE Education project is funded by The Medical Research Future Foundation Genomic Health Futures Mission (APP2015946) and will be carried out according to the National Statement on Ethical Conduct in Human Research (2007). The ethical aspects of this research project have been approved by the HREC of QIMR Berghofer, Project ID P3920.

**Terms used in this survey:**

Functional evidence - results from experimental assays, ranging from low throughput assays of a handful of variants, to extremely high throughput including MAVEs.

Variant Curation - process used to collate and review multiple different evidence types towards assigning variant pathogenicity (also known as variant classification). Includes review of suitability and weight of individual evidence types for a given variant/gene-disease relationship.

Variant Classification - assigning variant into one of multiple tiers to designate variant relationship to disease predisposition/causation e.g. Benign, Likely Benign, Variant of uncertain significance, Likely Pathogenic, Pathogenic, and additional categories depending on application.

We are seeking views from a wide range of stakeholders, so regardless of your background or training, your responses are valuable to us. All responses are anonymous. Summarised results will be used to support genetic variant interpretation education resources, and relevant findings may be published in a non-identifiable manner in peer reviewed scientific journals or relevant educational materials. Please note that survey data must be maintained indefinitely at QIMRB to comply with applicable legislations.

**Consent**

Your participation in this survey is entirely voluntary. You can choose not to participate at any point in the survey by navigating away from the page. Please be aware that by clicking submit at the end of this survey you will consent to the use of the survey summary results, as described above.

Please note that you may request further information or a copy of the study's anonymous summary results by entering your contact when requested at the completion of the survey; this information will not be linked in any way to your survey responses. If you have any concerns regarding this survey, please contact the project co-ordinator.

This survey should take around 20 minutes to complete.

**Please note we only require one response per participant. If you have previously completed this survey, you can exit this survey by simply navigating away from this page.**

**In your professional role, which of the following activities do you perform now, or anticipate you will perform in the future?**

| | do not perform | perform now | will perform in the future |
|---|---|---|---|
| **Variant classification for determining variant pathogenicity in a diagnostic setting** | ○ | ○ | ○ |
| **Variant classification for research purposes** | ○ | ○ | ○ |

| | | | |
|---|---|---|---|
| **Returning genetic test results to patients as part of diagnostic process** | ○ | ○ | ○ |
| **Discussing genetic test results with patients (but I am not responsible for returning the clinical diagnosis)** | ○ | ○ | ○ |
| **Participation in multidisciplinary meetings (Multidisciplinary Team Discussions, MTBs, or Molecular Tumour Boards, MTBs)** | ○ | ○ | ○ |
| **Participation in a ClinGen expert panel, including a Gene Curation Expert Panel (GCEP) or a Variant Curation Expert Panel (VCEP)** | ○ | ○ | ○ |

**In your professional role, which of the following functional evidence related activities do you perform now or anticipate you will perform in the future?**

| | do not perform | perform now | will perform in the future |
|---|---|---|---|
| **Evaluate functional data from a publicly available datasource or publication for research purposes** | ○ | ○ | ○ |
| **Curate functional evidence from a publicly available datasource or publication for the purpose of clinical variant classification** | ○ | ○ | ○ |
| **Use functional evidence as a catalyst to reassess a variant classification in the clinical setting** | ○ | ○ | ○ |
| **Request functional evidence from a diagnostic accredited laboratory to inform clinical interpretation of a variant** | ○ | ○ | ○ |
| **Request functional evidence from a research laboratory to inform clinical interpretation of a variant** | ○ | ○ | ○ |

**Please list any additional functional evidence related activities that you perform now or may perform in the future.**

**How do you rate your confidence in using the following categories of functional evidence for the purpose of genetic variant classification, on a scale of 1 (not at all comfortable) to 5 (very comfortable), OR NA - I don't interpret variants ?**

| | not at all comfortable | not comfortable | somewhere in the middle | comfortable | very comfortable |
|---|---|---|---|---|---|
| **Biochemical assays, for e.g. enzyme assays** | ○ | ○ | ○ | ○ | ○ |
| **Transcript assays, for e.g. splicing assays or transcriptome data** | ○ | ○ | ○ | ○ | ○ |
| **Cell models, for e.g. in vitro cell assay for cell survival** | ○ | ○ | ○ | ○ | ○ |
| **Animal models, for e.g. mouse model of a genetic disease** | ○ | ○ | ○ | ○ | ○ |
| **High throughput functional assays, including MAVEs** | ○ | ○ | ○ | ○ | ○ |

**Are there any functional evidence categories not listed above that you would consider using in genetic variant classification?**

**Do you follow a policy or procedure that specifies how to incorporate functional evidence into genetic variant classification?**

- ☐ No
- ☐ Yes - ACMG/AMP guidelines as per Richards et al 2015.
- ☐ Yes - ACMG/AMP guidelines and Brnich et al 2020 guidelines for functional evidence
- ☐ Yes - Internal SOP
- ☐ Yes - Other
- ☐ I do not know

**What resources do you use to source functional evidence for genetic variant classification?**

Are you aware of and do you use the following functional evidence approaches/resources?

| | do not know of this | aware of | use |
|---|---|---|---|
| **Brnich et al. 2020 (Functional evidence recommendations)** | ○ | ○ | ○ |
| **Walker et al. 2023 (Splicing recommendations)** | ○ | ○ | ○ |
| **MaveDB (Database of high throughput experimental data)** | ○ | ○ | ○ |
| **The Atlas of Variant Effects (AVE) Resources Page** | ○ | ○ | ○ |
| **ClinGen SVI functional assay assessment worksheet** | ○ | ○ | ○ |
| **VCEP specific guidance on functional evidence use** | ○ | ○ | ○ |
| **Ensembl VEP - MaveDB annotation** | ○ | ○ | ○ |

**How would you deal with conflicting evidence or results when using functional evidence for variant classification?**

**Which of the following might prevent your use of functional evidence in variant classification?**

- ☐ Lack of time
- ☐ Insufficient training
- ☐ Outside the scope of my role
- ☐ Cannot find functional data
- ☐ Lack of familiarity with specific assay
- ☐ The assay is too general (not gene-disease specific enough)
- ☐ Other

**What would improve your interaction with and use of functional evidence (existing or not)?**

- ☐ Education/training in functional evidence use
- ☐ Additional guidelines on functional evidence use
- ☐ Improved methods to source functional evidence
- ☐ Increased information on published functional data
- ☐ Improved access to commission creation of functional data
- ☐ Improved curation resourcing (more time/funding)

**What else might improve your interaction with and use of functional evidence?**

**Where such data exist for a variant/gene of interest, how likely are you to use high throughput functional data (including MAVE data) in clinical classification of a genetic variant?**

not at all likely — I would consider — very likely

Change the slider above to set a response

reset

**What has been your interaction with MaveDB (Multiplexed Assays of Variant Effect Database)?**

- ○ I do not know what MaveDB is
- ○ I know what MaveDB is but have not used it
- ○ I have accessed data via MaveDB
- ○ I incorporate data from MaveDB into variant classification

**Would you be interested in training/further education and/or additional resources to support your use of functional evidence?**

○ Yes ○ No

**Which of the following training and/or additional resources would you use to increase/improve your use/understanding of applying functional evidence? (please select all that apply)**

- ☐ Process guidelines
- ☐ Expert guidance documents (for e.g. from Variant Curation Expert Panels)
- ☐ Peer reviewed recommendations
- ☐ Tools to assist in functional assay evaluation
- ☐ Professional development opportunities
- ☐ Online learning resources
- ☐ I would not access
- ☐ Other?

**If you selected other, please list resources you would use to improve your use/understanding of functional evidence.**

**What aspects of functional evidence use would you like more understanding of?**

**What is your primary professional position?**

**How many years of professional experience do you have in your primary position?**

**Join the contact list: Add your preferred contact if you would like to receive the summary results of this survey and/or know more about functional evidence and the MAVE Education project**

Submit

Save & Return Later

Powered by REDCap
